# Asymptomatic *Plasmodium falciparum* Carriage at the End of the Dry Season is Associated with Subsequent Infection and Clinical Malaria in Eastern Gambia

**DOI:** 10.1101/2023.09.29.23296347

**Authors:** Balotin Fogang, Lionel Lellouche, Sukai Ceesay, Sainabou Drammeh, Fatou K. Jaiteh, Marc-Antoine Guery, Jordi Landier, Cynthia Haanappel, Janeri Froberg, David Conway, Umberto D’Alessandro, Teun Bousema, Antoine Claessens

**Affiliations:** LPHI, MIVEGEC, University of Montpellier, CNRS, INSERM, Montpellier, France; Medical Research Council Unit The Gambia at the London School of Hygiene and Tropical Medicine, Banjul, The Gambia; Aix Marseille Univ, IRD, INSERM, SESSTIM, ISSPAM, 27 boulevard Jean Moulin, 13005, Marseille, France; Radboud university medical center, Radboud Institute for Health Sciences, Department of Medical Microbiology, Nijmegen, The Netherlands; Department of Infection Biology, London School of Hygiene and Tropical Medicine, London, UK

**Keywords:** *Plasmodium falciparum*, asymptomatic, clinical malaria, seasonal transmission

## Abstract

**Background:** Chronic carriage of asymptomatic low-density *Plasmodium falciparum* parasitaemia in the dry season may support maintenance of acquired immunity that protects against clinical malaria. However, the relationship between chronic low-density infections and subsequent risk of clinical malaria episodes remains unclear.

**Methods:** In a 2-years study (December 2014 to December 2016) in eastern Gambia, nine cross-sectional surveys using molecular parasite detection were performed in the dry and wet season. During the 2016 malaria transmission season, passive case detection identified episodes of clinical malaria.

**Results:** Among the 5,256 samples collected, 444 (8.4%) were positive for *P. falciparum*. A multivariate model identified village of residence, male sex, age ≥5 years old, anaemia, and fever as independent factors associated with *P. falciparum* parasite carriage. Infections did not cluster over time within the same households or recurred among neighbouring households. Asymptomatic parasite carriage at the end of dry season was associated with a higher risk of infection (Hazard Ratio, HR= 3.0, p< 0.0001) and clinical malaria (HR= 1.561, p= 0.057) during the following transmission season. Age and village of residence were additional predictors of infection and clinical malaria during the transmission season.

**Conclusion:** Chronic parasite carriage during the dry season is associated with an increased risk of malaria infection and clinical malaria. It is unclear whether this is due to environmental exposure or to other factors.

## Introduction

Malaria continues to be a significant public health challenge in several parts of the world, where it is responsible for a high burden of morbidity and mortality [1,2]. Despite the impact of standard control interventions, asymptomatic *P. falciparum* infections continue to pose a challenge to control efforts as they maintain residual transmission [3]. Where malaria transmission is seasonal, most cases of clinical malaria occur between July and December, a period covering the seasonal rains and the 2 months immediately following them, with a peak in cases in November, while the dry season (January-June) is a period of low transmission, with mainly asymptomatic chronic infections acquired during the previous transmission season [4,5].

As malaria transmission has declined considerably in The Gambia [1], thanks to wide scale up of vector control interventions, the prevalence of infection has become increasingly heterogeneous [6–8]. To sustain the decline in malaria transmission and move towards elimination, there is a need to evaluate and improve existing community-based interventions through ongoing epidemiological surveillance and identification of associated factors. A large proportion of asymptomatic individuals at the end of the transmission season remains infected for several months, from January to June [9]. However, the relationship between these chronic infections and the risk of infection and clinical malaria during the following transmission season remains unclear, with conflicting information; some studies report a protective effect [10–13] while others a higher risk of clinical malaria [14–17] or no consistent association [6,18]. Such differences may be influenced by the age of the participants [17,18], the intensity of malaria transmission [6,18] and the number of circulating *P. falciparum* clones [13,14,19]. Importantly, variation in parasite exposure may also play a role.

Here, to characterise the dynamics of malaria transmission and identify associated risk factors, we conducted a total of 9 cross-sectional surveys during the dry and wet seasons, starting in December 2014. From July 2016, a longitudinal follow-up of treated asymptomatic and non-infected participants with passive *P. falciparum* detection was carried out to assess the impact of asymptomatic parasitaemia on subsequent infection and clinical malaria during the transmission season.

## Materials and methods

### Ethical considerations

The study protocol was reviewed and approved by the Gambia Government/MRC Joint Ethics Committee (SCC 1476, SCC 1318, L2015.50) and by the London School of Hygiene & Tropical Medicine ethics committee (Ref 10982). The field studies were also approved by local administrative representatives, the village chiefs. Written informed consent was obtained from participants over 18 years old and from parents/guardians for participants under 18 years. Written assent was obtained from all individuals aged 12-17 years.

### Study design and population

The study was carried out in four villages (Madina Samako: K, Njayel: J, Sendebu: P, and Karandaba: N) in Upper River Region (URR), eastern Gambia (Figure 1A) [9,20]. Over a two-year period, two cohorts of individuals at least two years old were recruited; one cohort was followed up between December 2014 and March 2016, and the other between July and December 2016 (Figure 1B). All households were invited to participate in the study. The number of participants per household ranged from 1 to 79, with a median of 7 (Q1, Q3 4, 11). Five surveys covering the end of the transmission season (December 2014 and November and December 2015) and the following dry season (April 2015 and March 2016) were carried out in the first cohort in two-villages (J and K) (Figure 1B). Surveys on the second cohort were carried out during the 2016 transmission season (July, October, November, and December) in the four study villages (Figure 1B). During the surveys, structured questionnaires were used to record individual information (village, household, age, sex, bed net use, history of fever, temperature) from all participants. Moreover, households in the villages J and K were geolocated using a global positioning system (GPS). During the 2016 transmission season (July-December), a resident nurse delivered complimentary basic healthcare to all participants. Suspected malaria cases (axillary temperature > 37.5 °C) were tested with a Rapid Diagnostic Test (CareStart) and, if positive, treated with artemether-lumefantrine.

**Figure 1.**
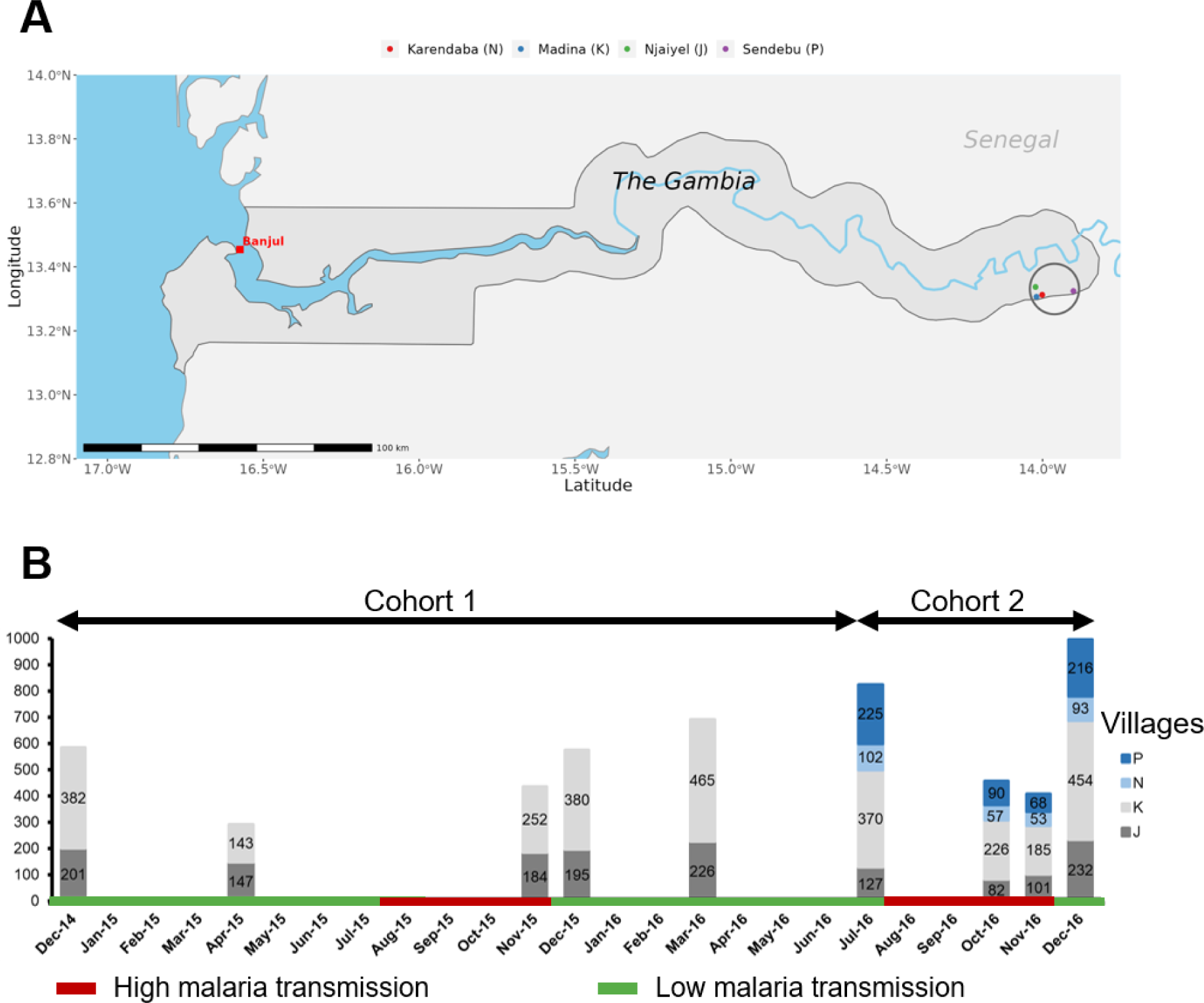
Study site and design. A) Map of The Gambia with the study site (4 villages) circled. B) Presentation of the two cohorts and the 2-year sampling period in the four villages. The bar charts show the cumulative number of fingerprick samples per timepoint for each village. The green colour represents the low malaria transmission season and the red colour represents the high transmission malaria season in The Gambia. The colours in the bars indicate village of residence (P= blue, N= light blue, K= light grey, J= grey).

### Sample collection and malaria diagnostic

At each survey, approximately 200µl of blood was collected by fingerprick into an EDTA-coated tube. A few drops were used for a thick blood film and dry blood spots on filter paper (Whatman 3 Corporation, Florham Park, NJ, USA). Haemoglobin (Hb) was measured by HaemoCue 301 machine. Blood samples from EDTA tubes were processed on the same day for leucodepletion using a cellulose-based column (MN2100ff, similar to CF11) [21].

Thick blood films were stained with a 10% Giemsa solution for 10 minutes and parasite density estimated by counting the number of parasites per at least 200 white blood cells (WBC) and assuming 8,000 WBC/μL. Slides were considered negative if no parasites were detected by two microscopists after reading 500 WBC. A 20 % error check was used to identify discrepancies between slide readers. All discordant results were read by a senior microscopist whose results were taken as final.

For samples collected between December 2014 and March 2016, DNA extracted from leukocyte-depleted red blood cells using the QIAgen MiniPrep kit was analyzed by nested PCR targeting *P. falciparum* 18S small subunit rRNA (ssRNA) genes [22].

For samples collected from July to December 2016, DNA was extracted from dry blood spot (DBS) by Chelex method [23] and analyzed by 18S small subunit of ribosomal RNA (rRNA) qPCR and *var* gene acid terminal sequence (varATS) qPCR [24], as detailed in Supplementary Figure 1.

### Statistical analysis

Data were collected on paper-based case report forms (CRFs), double entered, and checked for errors and discrepancies before statistical analysis. Statistical analyses were performed using R studio version 4.2.3 [25]. Age was categorized into three groups: < 5 years, 5–14 years and ≥ 15 years. Anaemia was classified as mild, moderate or severe [26] (**Supplementary Table 1)**. Comparisons of proportions between groups were performed using the chi-square χ2 test; Spearman’s correlation test was used to test the association between quantitative variables. 95% confidence intervals for prevalence were calculated using standard error based on Wilson Score Interval.

A multivariate mixed-effects logistic regression model, implemented with the R lme4 package, was used to determine factors independently associated with qPCR-detected *P. falciparum* infection. This modelling approach incorporates both fixed and random effects, making it suitable for analysing data with repeated or clustered observations. The mixed-effects logistic regression model assumes that observations within groups or clusters defined by random effects must be independent, and there should be no perfect separation in the data. In this model all fixed-effect variables were adjusted for clustering at different months of data collection (time of sampling), at the household level, and at the individual level, with the individual level nested within the household level. This adjustment helps mitigate potential biases stemming from temporal and household-level variability. The predictive factors (fixed-effects) included villages, age, sex, anaemia, history of fever in the past week and fever at inclusion (temperature> 37.5°C). For this analysis, only individuals in the second cohort were included as complete epidemiological information for the first cohort was not available.

To assess the risk factors associated with subsequent infection or the first clinical malaria episode during the transmission season, both univariate and multivariate shared frailty mixed-effects survival models were applied These models were implemented using the R survival and coxme packages. They integrate components of survival analysis and mixed-effects modelling, taking into consideration the inherent clustering or interdependence of observations within groups or clusters. The explained variables in separated models included the occurrence of infection or clinical malaria during the transmission season. In these models, individuals were clustered according to the timing of reinfection or the onset of clinical malaria, with each interval spanning one month. The random effect in the model pertained to households, and the covariates included the infection status at the end of dry season, villages, age, sex, anaemia status, history of fever in the past week, fever at inclusion (temperature> 37.5°C). Kaplan-Meier survival curves were used to visualize the progression of malaria incidence or clinical cases during the transmission season.

Spatial analysis of clusters of *Plasmodium* infections (hotspot) was performed using the Bernoulli method in SaTScan version 10.1.2 [27]. In all analyses, P values < 0.05 were considered statistically significant. All the graphs were performed using R studio.

## Results

### Characteristics of the study cohorts

Over a period of 2 years, 5256 blood samples were collected from 1471 individuals aged 2 to 85 years (Fig. 1). The first cohort included 1003 participants and the second cohort 1284 participants, with 789 of them in both cohorts (Table 1). 46.3% (458) of cohort 1 were males and the median age was 11 years (Q1, Q3 5.5, 27). In the cohort 2, the median age was 12 years (Q1, Q3 6, 29), with more females than males. Anaemia prevalence was 42.8%, mostly mild (22.5%) and moderate (18.9%), with few cases of severe anaemia (1.4%) (Table 1). Fever at inclusion was rare (0.8%). Bed net use was high (76.4%).

**Table 1:** Characteristics of the study population.

| Characteristics |  | Cohort 1: December 2014 –<br>March 2016, % (n/N) | Cohort 2: July 2016 –<br>December 2016, % (n/N) |
| --- | --- | --- | --- |
| Male sex |  | 46.3 (458/989) | 44.1 (565/1280) |
| Villages |  |  |  |
|  | J | 33.4 (335/1003) | 20.6 (264/1284) |
|  | K | 66.6 (668/1003) | 48.4 (622/1284) |
|  | N | - | 11.1 (142/1284) |
|  | P | - | 19.9 (256/1284) |
| Age groups (years) |  |  |  |
|  | < 5 | 22.1 (219/1003) | 16.2 (207/1280) |
|  | 5-14 | 33.9 (335/1003) | 39.3 (503/1280) |
|  | >15 | 44.0 (435/1003) | 44.5 (570/1280) |
| Anaemia* |  |  |  |
|  | Non-anaemic | - | 57.2 (688/1203) |
|  | Mild | - | 22.5 (271/1203) |
|  | Moderate | - | 18.9 (227/1203) |
|  | Severe | - | 1.4 (17/1203) |
| Slept under a LLIN last night |  | - | 76.4 (916/1195) |
| History of fever in the past week |  | - | 4.0 (48/1195) |
| Fever at inclusion |  | - | 0.8 (10/860) |

\*Anaemia is classified based on age and sex (WHO, 2011). LLIN: Long-lasting insecticidal net; - = not determined

### *Plasmodium falciparum* prevalence in cohort 1

Between December 2014 and March 2016, 155 (6%) of 2575 samples collected were positive. Prevalence varied substantially over time, between 1.7% and 5.4% during the dry season and up to 14.4% during the transmission season (Supplementary Figure 2). Children under 5 years of age were less likely to be infected (Supplementary Figure 2), possibly reflecting the effects of Seasonal Malaria Chemoprevention (SMC). Parasite prevalence in older children and adults was similar and peaked towards the end of the transmission season (Supplementary Figure 2).

### VarATS qPCR is more sensitive than 18S qPCR and microscopy

To increase the sensitivity of the *P. falciparum* detection relative to microscopy, two quantitative PCR approaches targeting the 18S locus of the conserved small subunit of ribosomal RNA (18S rRNA, 5-8 copies/genomes) and the acidic terminal sequence of the var gene (varATS, ∼60 copies/genomes) were evaluated using a set of samples from cohort 2. Using serial dilutions of the laboratory-adapted *P. falciparum* 3D7 strain to determine analytical sensitivity, varATS reached the limit of detection (LOD) of 0.22 parasites/µL of blood and was around 10 times more sensitive than standard 18S rRNA qPCR with the LOD of 2.2 parasites/µL (Supplementary Figure 1). Out of 1202 fingerprick samples from this cohort screened by the 3 techniques, 11.7% (141), 10.1% (121) and 3.9% (47) samples were positive for *P. falciparum* using varATS, 18S rRNA qPCR and light microscopy, respectively (Figure 2A). Of 166 *Plasmodium* infections detected by the three diagnostic tests combined, 18.9% (35) were only detected by varATS, while 10.2% (17) (p= 0.0066) and 4.2% (7) (p< 0.0001) were detected only by 18S rRNA qPCR and microscopy, respectively (Figure 2B and Supplementary Figure 3). There was a high level of agreement between varATS and 18S rRNA qPCR, with 95.3% (1146 varATS+/18SRNA+ or varATS-/18SRNA-) samples with the same results for both methods (Supplementary Figure 3). Parasite densities determined by varATS and 18S rRNA qPCR correlated strongly with parasitemia quantified by microscopy (r= 0.76, P< 0.0001 and r= 0.86, P< 0.0001 for VarATS and 18S rRNA qPCR, respectively), as well as with each other (r= 0.94, P< 0.0001) (Figure 2C). Because of the high concordance between the two molecular methods varATS and 18S rRNA qPCR, results from the two molecular *Plasmodium* detection tools were combined in a single analysis.

**Figure 2.**
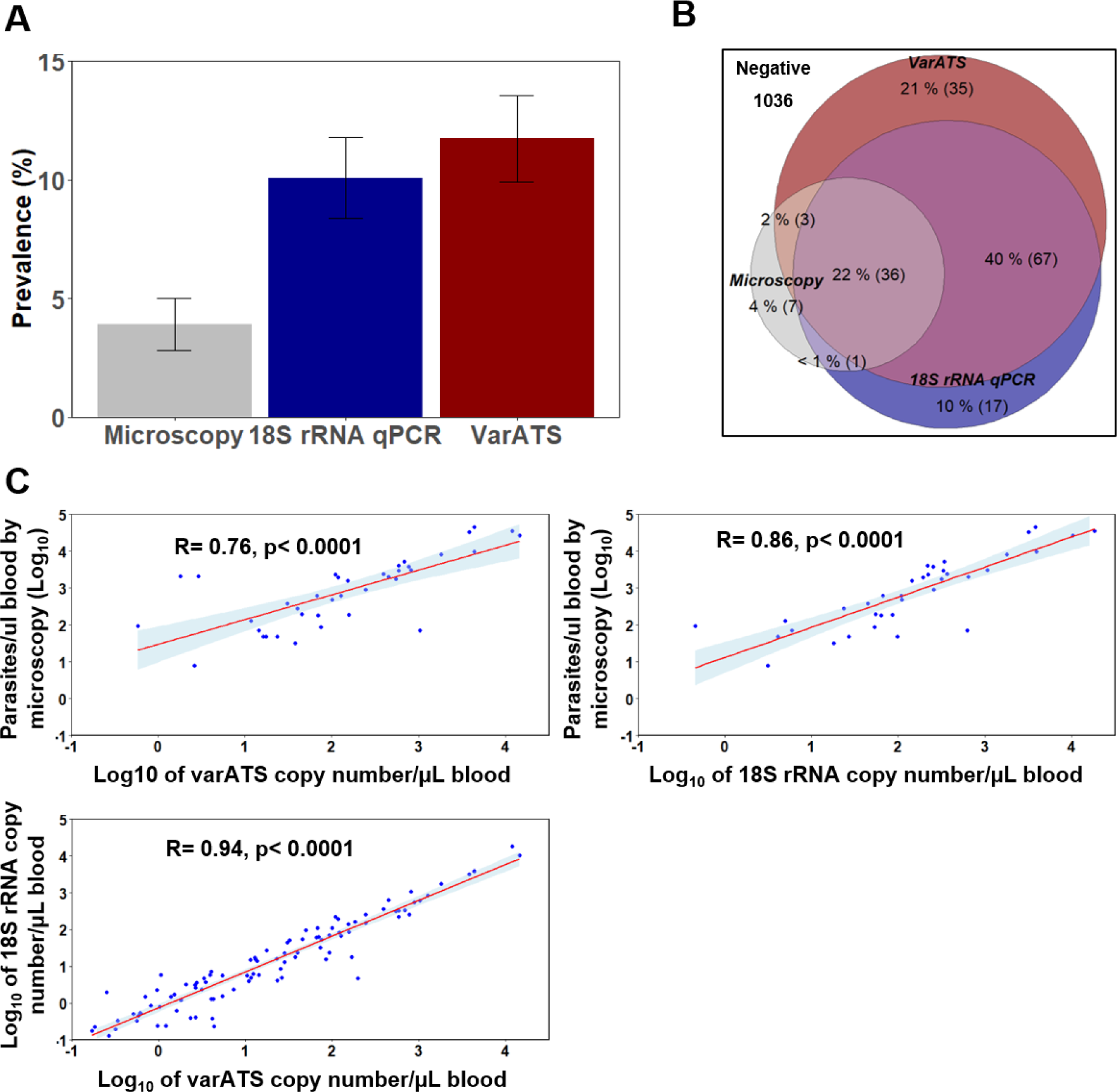
*Plasmodium* detection by different diagnostic tools. A) Proportion of *P. falciparum* detected with different diagnostic methods. B) Venn diagram of *Plasmodium* positivity by varATS, 18S rRNA qPCR and/or microscopy. C) Correlation in parasitaemia between diagnostic tools. Error bars in Fig A correspond to 95% confidence interval for estimated prevalence. Spearman rank correlation (panel C) is presented as the best fit line and the coefficient (r), as well as the p-value (p), are shown for each correlation pair between parasitaemia. LM: light microscopy. qPCR: quantitative polymerase chain reaction. rRNA: Ribosomal ribonucleic acid. ATS: acidic terminal segment.

### *Plasmodium falciparum* prevalence in cohort 2

*P. falciparum* prevalence was 9.3% (77/824) in July, 9.7% (44/455) in October, 20.6% (84/407) in November, and 12.9% (128/995) in December. This trend was observed in all age groups (Figure 3A). As shown in Figure 3A, children under 5 years of age had the lowest prevalence while older children the highest prevalence (Figure 3A). Bednet usage was high in all age groups (81% in < 5, 74% in 5-14, 77% in ≥ 15 years). Median varATS parasitaemia was usually higher in the older children than adults and young children (5-14 vs <5 years, p= 0.1553; 5-14 vs ≥15 years, p< 0.0001), but not in all surveys (Figure 3B).

**Figure 3.**
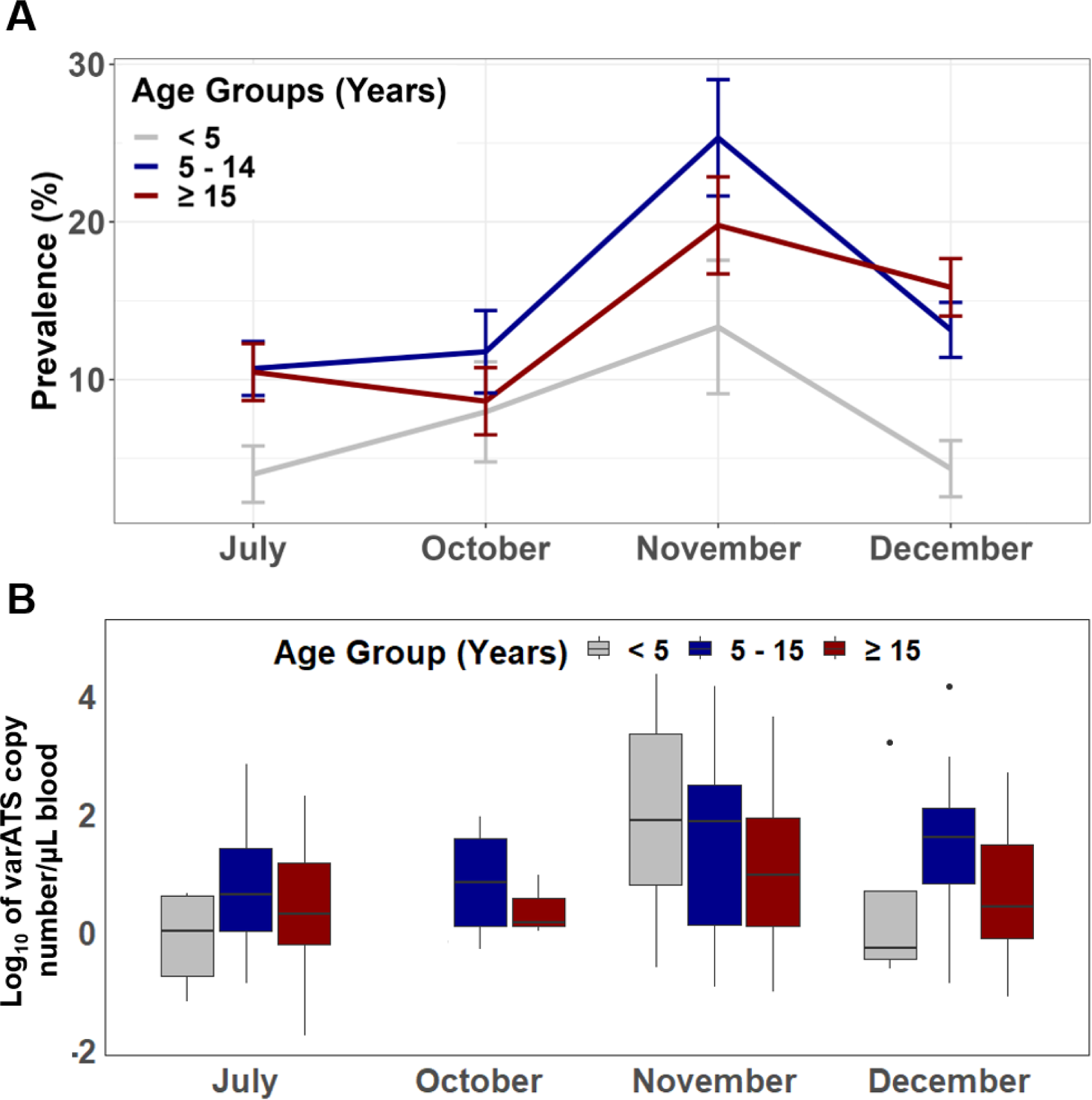
Trend analysis for *P. falciparum* infection prevalence (A) and parasitaemia (B) by age groups in the second cohort (2016). In A, error bars represent 95% confidence intervals for prevalence.. Box in B plots representing interquartile ranges (25th, median and 75th percentile, whisker percentile (1st and 99th)). (B) Parasite density by varATS was not available for October in the <5 year group.

### Risk factors associated with *P. falciparum* infection

A multivariate mixed-effects logistic regression model shows that sex, age, anaemia, fever at inclusion, and village of residence were independent factors associated with *P. falciparum* infections (Table 2). Indeed, risk of infection was 3-fold higher (OR= 3.134, p< 0.0001) in older children and 2.5-fold higher (OR= 2.616, p< 0.0001) in adults than in young children, likely indicating the effect of SMC in this age group.

**Table 2:** Assessment of associated risk factors of *P. falciparum* infection.

| Factors | Description (n) | % infections (n) | OR [95% CI] | P value |
| --- | --- | --- | --- | --- |
| Sex | F (1513) | 11.4 (172) | Ref |  |
|  | **M (1162)** | **13.9 (162)** | **1.336 [1.031-1.731]** | **0.0280** |
| Age groups (Years) | < 5 (474) | 6.33 (30) | Ref |  |
|  | **5-14 (1078)** | **13.8 (149)** | **3.134 [2.041-4.813]** | **< 0.0001** |
|  | **≥ 15 (1123)** | **13.7 (154)** | **2.616 [1.697-4.032]** | **< 0.0001** |
| Village of residence | J (542) | 10.3 (56) | 1.616 [0.895-2.916] | 0.1110 |
|  | **K (1235)** | **11.9 (147)** | **1.848 [1.085-3.146]** | **0.0236** |
|  | N (305) | 7.9 (24) | Ref |  |
|  | **P (599)** | **17.7 (106)** | **3.499 [2.026-6.043]** | **< 0.0001** |
| History of fever in the past week | Yes (131) | 14.5 (19) | 1.021 [0.590-1.767] | 0.9382 |
|  | No (2306) | 12.49 (288) | Ref |  |
| Anaemia* | Non-anaemic (1390) | 10.6 (147) | Ref |  |
|  | **Mild (550)** | **13.6 (75)** | **1.577 [1.155-2.155]** | **0.0042** |
|  | **Moderate (493)** | **16.2 (80)** | **1.759 [1.294-2.392]** | **0.0003** |
|  | **Severe (37)** | **29.7 (11)** | **3.942 [1.818-8.548]** | **0.0005** |
| Fever at inclusion | **Yes (19)** | **31.6 (6)** | **4.374 [1.496-12.78]** | **0.0070** |
|  | No (2446) | 12.6 (307) | Ref |  |

Only individuals from the second cohort (N= 2681) were included in this multivariate mixed-effects logistic regression model because complete epidemiological information were only available for these individuals. In this model all fixed-effect variables were adjusted for clustering at different months of data collection (time of sampling), at the household level, and at the individual level, with the individual level nested within the household level. \*Anaemia determined by age and sex according to the OMS 2011 report (OMS, 2011). Significant estimates are bolded. OR: odds ratio. CI: Confident interval. Ref: reference.

Residents of villages K and P were more at risk of infection than those of village N (OR= 1.848, p= 0.0236 and OR= 3.499, p< 0.0001 for villages K and P, respectively). Individuals with different degree of anaemia had higher odds of being infected by *P. falciparum* compared to non-anaemic individuals (Table 2). Fever at inclusion was strongly associated with infection (OR= **4.374**, p= 0.007).

As malaria prevalence varied substantially between and within villages, the analysis investigated whether some households were hotspots of malaria transmission. Analyses focused on villages J and K, as data are available over a 2-year study period. By utilizing the Bernoulli spatial scan statistic in Satscan algorithm, two clusters in village J with a high prevalence of *P. falciparum* infections were identified (Supplementary Table 2). Similarly, in village K, three clusters of high *Plasmodium falciparum* infections at two different times (November and December 2016) were identified (P< 0.05) (Supplementary Table 2). However, when *P. falciparum* prevalence at the household level in the two villages was plotted over time, no evidence of clustering of infection over time within the same households or recurrent infections mong neighbouring households was found (Supplementary Figure 4). This suggests that transmission is stochastic, with no evidence of sustained malaria hotspots in these villages.

### Asymptomatic parasitaemia at the end of dry season is associated with infection and clinical malaria during the following transmission season

The individual risk of being infected during the transmission season according to the infection status at the beginning of the transmission season was investigated (Figure 4 and Table 3). In July 2016, 67 individuals were asymptomatic carriers and treated with artesunate-lumefantrine antimalarials; 68.7% (46) of them were re-infected during the transmission season. Among the 613 individuals non-infected in July 2016, 28.5% (175) of them became infected during the same period (HR= 3.338, p< 0.0001) (Figure 4 and Table 3). The association between infection status and risk of infection during the transmission season remains significant after adjustment for age and village (HR= 3.0, p< 0.0001) (Table 3). Multivariate analysis shows that age and village of residence were independently associated with risk of infection (HR= 2.767, p< 0.0001 for age 5-14 years, HR= 3.063, p< 0.0001 for age ≥ 15 years and HR= 2.375, p= 0.0012 for village P).

**Figure 4.**
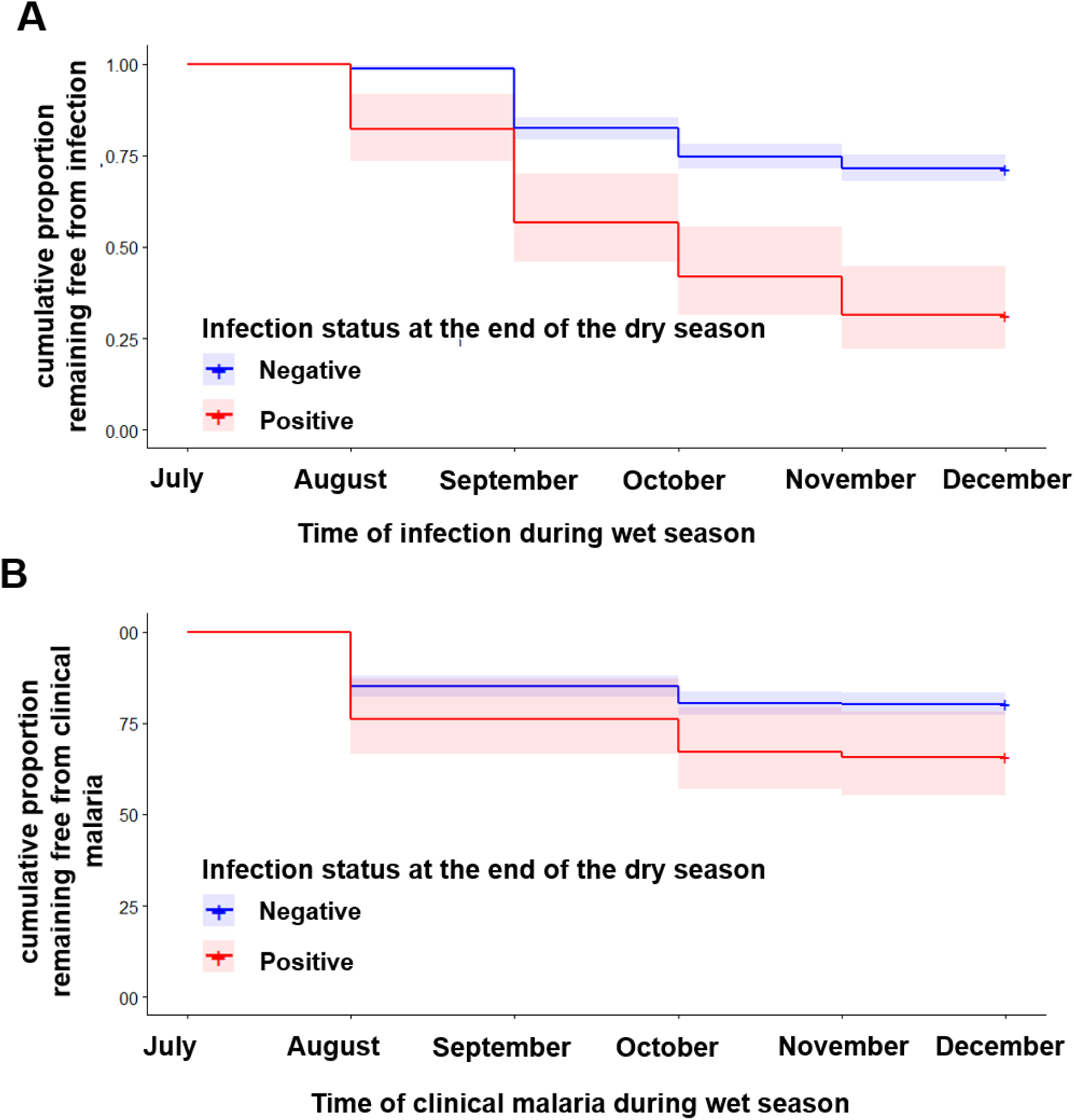
Asymptomatic *P. falciparum* carriage at the end of dry season and during the subsequent transmission season. Being carrier in the dry season increases the risk of both *P. falciparum* infection (A) and malaria symptoms in the following transmission season (B). Time to infection and/or onset of clinical malaria were grouped into a one-month interval.

**Table 3:**
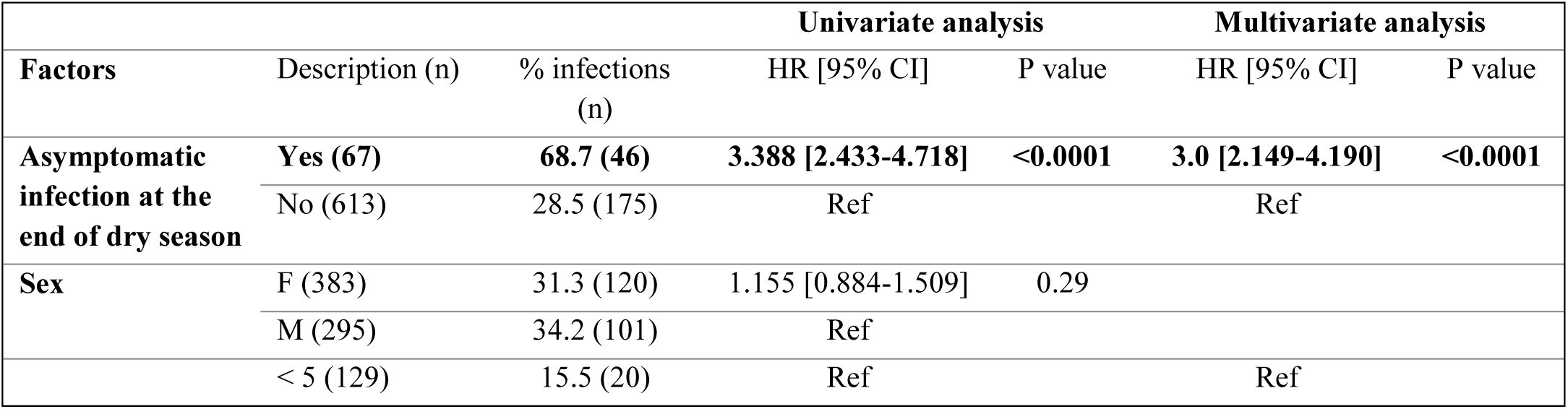
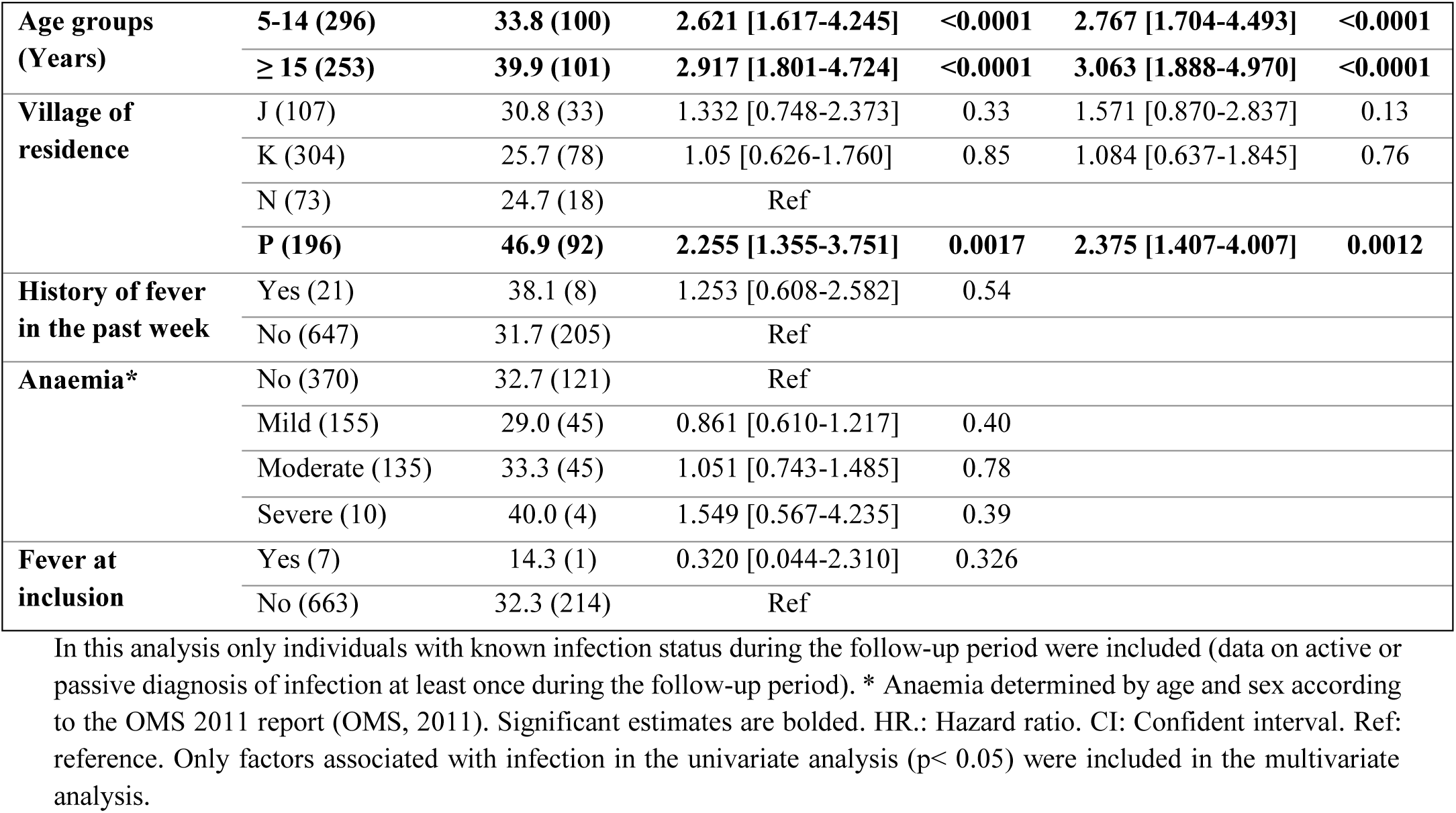
Predicted hazard of *P. falciparum* infection during the malaria transmission season.

| Factors | Description (n) | Univariate analysis |  |  | Multivariate analysis |  |
| --- | --- | --- | --- | --- | --- | --- |
|  |  | % infections (n) | HR [95% CI] | P value | HR [95% CI] | P value |
| Asymptomatic infection at the end of dry season | Yes (67) | 68.7 (46) | 3.388 [2.433-4.718] | <0.0001 | 3.0 [2.149-4.190] | <0.0001 |
|  | No (613) | 28.5 (175) | Ref |  | Ref |  |
| Sex | F (383) | 31.3 (120) | 1.155 [0.884-1.509] | 0.29 |  |  |
|  | M (295) | 34.2 (101) | Ref |  |  |  |
|  | < 5 (129) | 15.5 (20) | Ref |  | Ref |  |
| **Age groups (Years)** | **5-14 (296)** | **33.8 (100)** | **2.621 [1.617-4.245]** | **<0.0001** | **2.767 [1.704-4.493]** | **<0.0001** |
|  | **≥ 15 (253)** | **39.9 (101)** | **2.917 [1.801-4.724]** | **<0.0001** | **3.063 [1.888-4.970]** | **<0.0001** |
| **Village of residence** | J (107) | 30.8 (33) | 1.332 [0.748-2.373] | 0.33 | 1.571 [0.870-2.837] | 0.13 |
|  | K (304) | 25.7 (78) | 1.05 [0.626-1.760] | 0.85 | 1.084 [0.637-1.845] | 0.76 |
|  | N (73) | 24.7 (18) | Ref |  |  |  |
|  | **P (196)** | **46.9 (92)** | **2.255 [1.355-3.751]** | **0.0017** | **2.375 [1.407-4.007]** | **0.0012** |
| **History of fever in the past week** | Yes (21) | 38.1 (8) | 1.253 [0.608-2.582] | 0.54 |  |  |
|  | No (647) | 31.7 (205) | Ref |  |  |  |
| **Anaemia\*** | No (370) | 32.7 (121) | Ref |  |  |  |
|  | Mild (155) | 29.0 (45) | 0.861 [0.610-1.217] | 0.40 |  |  |
|  | Moderate (135) | 33.3 (45) | 1.051 [0.743-1.485] | 0.78 |  |  |
|  | Severe (10) | 40.0 (4) | 1.549 [0.567-4.235] | 0.39 |  |  |
| **Fever at inclusion** | Yes (7) | 14.3 (1) | 0.320 [0.044-2.310] | 0.326 |  |  |
|  | No (663) | 32.3 (214) | Ref |  |  |  |

In this analysis only individuals with known infection status during the follow-up period were included (data on active or passive diagnosis of infection at least once during the follow-up period). \* Anaemia determined by age and sex according to the OMS 2011 report (OMS, 2011). Significant estimates are bolded. HR.: Hazard ratio. CI: Confident interval. Ref: reference. Only factors associated with infection in the univariate analysis (p< 0.05) were included in the multivariate analysis.

Individuals with a malaria parasite infection in July 2016 had a 2-fold higher risk of experiencing a clinical malaria episode than those without infection (HR= 1.825, p= 0.0086); this effect persisted after adjusting for age, village and anaemia (HR= 1.561, p= 0.057) (Table 4). Additionally, age and villages were also independent predicting factors for clinical malaria during the transmission season (HR= 3.404, p=0.0003, HR= 3.951, p p< 0.0001 and HR= 2.717, p= 0.0054 for age 5-14 years, age ≥ 15 years and village P, respectively). Together, these data indicate that asymptomatic carriage of parasites during the dry season is associated with an increased risk of infection and clinical malaria during the following transmission season.

**Table 4:**
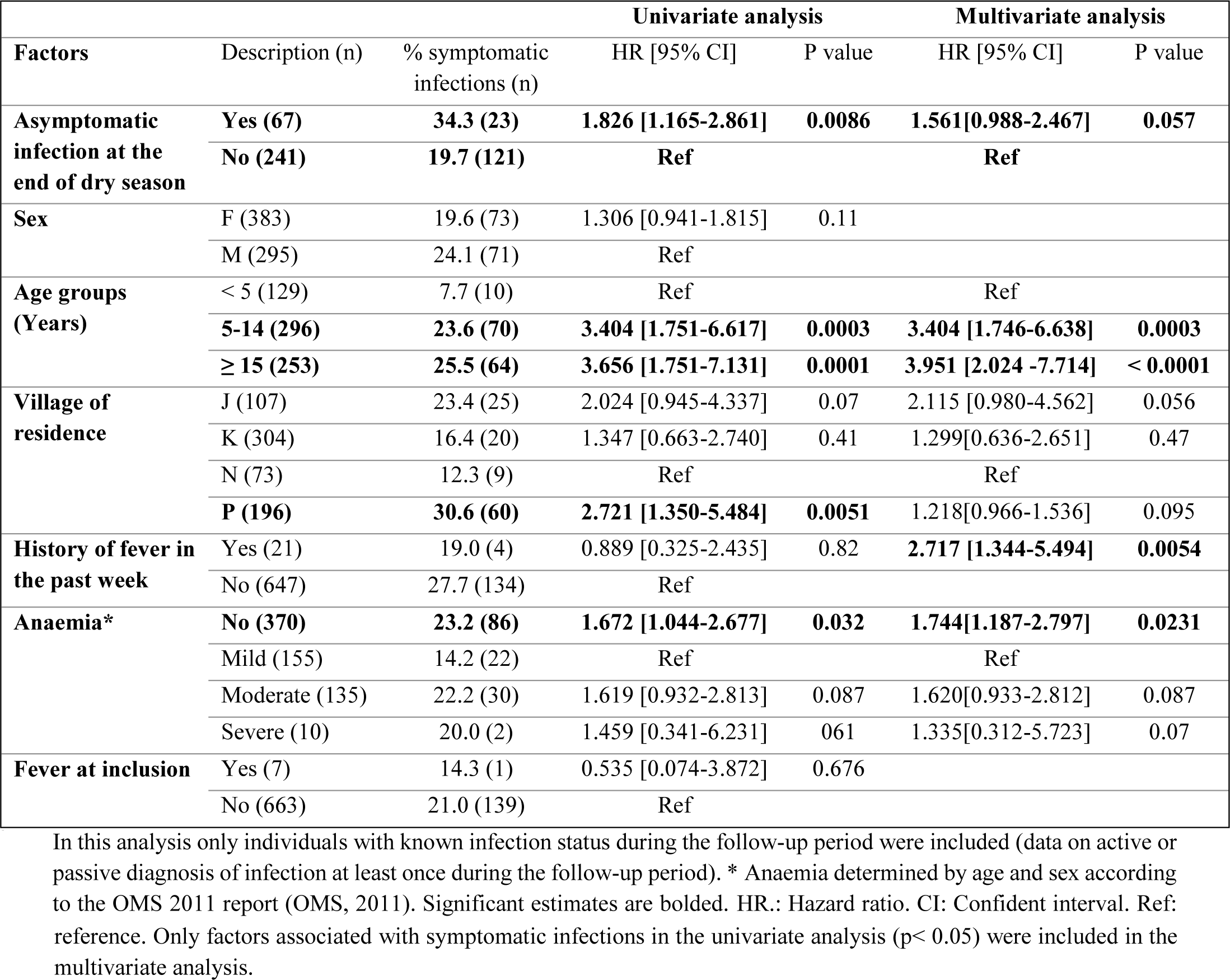
Predicted hazard of clinical malaria during the malaria transmission season.

| **Factors** | Description (n) | **Univariate analysis** |  |  | **Multivariate analysis** |  |
| --- | --- | --- | --- | --- | --- | --- |
|  |  | % symptomatic infections (n) | HR [95% CI] | P value | HR [95% CI] | P value |
| **Asymptomatic infection at the end of dry season** | **Yes (67)** | **34.3 (23)** | **1.826 [1.165-2.861]** | **0.0086** | **1.561[0.988-2.467]** | **0.057** |
|  | **No (241)** | **19.7 (121)** | **Ref** |  | **Ref** |  |
| **Sex** | F (383) | 19.6 (73) | 1.306 [0.941-1.815] | 0.11 |  |  |
|  | M (295) | 24.1 (71) | Ref |  |  |  |
| **Age groups (Years)** | < 5 (129) | 7.7 (10) | Ref |  | Ref |  |
|  | **5-14 (296)** | **23.6 (70)** | **3.404 [1.751-6.617]** | **0.0003** | **3.404 [1.746-6.638]** | **0.0003** |
|  | **≥ 15 (253)** | **25.5 (64)** | **3.656 [1.751-7.131]** | **0.0001** | **3.951 [2.024 -7.714]** | **< 0.0001** |
| **Village of residence** | J (107) | 23.4 (25) | 2.024 [0.945-4.337] | 0.07 | 2.115 [0.980-4.562] | 0.056 |
|  | K (304) | 16.4 (20) | 1.347 [0.663-2.740] | 0.41 | 1.299[0.636-2.651] | 0.47 |
|  | N (73) | 12.3 (9) | Ref |  | Ref |  |
|  | **P (196)** | **30.6 (60)** | **2.721 [1.350-5.484]** | **0.0051** | 1.218[0.966-1.536] | 0.095 |
| **History of fever in the past week** | Yes (21) | 19.0 (4) | 0.889 [0.325-2.435] | 0.82 | **2.717 [1.344-5.494]** | **0.0054** |
|  | No (647) | 27.7 (134) | Ref |  |  |  |
| **Anaemia\*** | **No (370)** | **23.2 (86)** | **1.672 [1.044-2.677]** | **0.032** | **1.744[1.187-2.797]** | **0.0231** |
|  | Mild (155) | 14.2 (22) | Ref |  | Ref |  |
|  | Moderate (135) | 22.2 (30) | 1.619 [0.932-2.813] | 0.087 | 1.620[0.933-2.812] | 0.087 |
|  | Severe (10) | 20.0 (2) | 1.459 [0.341-6.231] | 0.61 | 1.335[0.312-5.723] | 0.07 |
| **Fever at inclusion** | Yes (7) | 14.3 (1) | 0.535 [0.074-3.872] | 0.676 |  |  |
|  | No (663) | 21.0 (139) | Ref |  |  |  |

In this analysis only individuals with known infection status during the follow-up period were included (data on active or passive diagnosis of infection at least once during the follow-up period). \* Anaemia determined by age and sex according to the OMS 2011 report (OMS, 2011). Significant estimates are bolded. HR.: Hazard ratio. CI: Confident interval. Ref: reference.
reference. Only factors associated with symptomatic infections in the univariate analysis ( $p < 0.05$ ) were included in the multivariate analysis.

## Discussion

In The Gambia, there is a need to evaluate and improve existing community-based interventions through ongoing epidemiological surveillance and the identification of risk factor for parasite carriage. The dynamics and the factors associated with *P. falciparum* infections, and the link between asymptomatic carriage before the transmission season and risk of infection and clinical malaria were investigated over 2 years.

Consistent with previous studies in The Gambia [6,13,20], the prevalence of infection was lower in children under 5 years of age than in older children and adults, possibly due to the seasonal malaria chemoprophylaxis programme (monthly antimalarial treatment during the transmission season) [28,29]. The higher prevalence of infection and high parasite densities in older children than in adults may be the result of acquired immunity that makes adults better protected and able to better control parasite growth when infected [30]. Furthermore, in line with acquired immunity with age, when infected during the transmission season, children under 5 years old had a higher parasite density compared to adults. We observed that individuals with different degrees of anaemia were more likely to be infected with *P. falciparum* than non-anaemic individuals. This observation appears to be the consequence of *Plasmodium* infection rather than the cause of infection, given that malaria is known to be the main cause of anaemia in endemic areas [31,32].

There was significant variability in malaria prevalence between villages. These findings demonstrate the heterogeneity of malaria transmission in the eastern Gambia, even among neighbouring villages. Similarly, in a previous study [20], significant variation in malaria prevalence in this region was observed, ranging from 21.17% to 49.13%. This discrepancy was attributed not only to the observed higher insecticide resistance [33,34] but also to differences in vector composition. Indeed, *An. gambiae s.s.* is more common in villages away from The Gambia river, while *An. arabiensis* and *An. coluzzii* are more widespread near the river [34].

Due to the variation in malaria prevalence among neighbouring villages, there is a hypothesis regarding the presence of sustained malaria transmission hotspots within the villages, where certain households, based on the characteristics of the individuals and their environment, could sustain transmission in the area. Although some households exhibited significantly high malaria prevalence within villages, these households were not consistently the same, with no evidence of hotspot households spreading malaria, suggesting that the transmission pattern follows a stochastic pattern in the area [35]. Therefore, a targeted malaria prevention approach focusing households with highest prevalence is unlikely to yield a lasting effect in this area. However, treating all malaria cases identified in cohort 2 may have altered spatio-temporal patterns.

Asymptomatic carriage of *P. falciparum* at the end of the dry season was associated with both an increased risk of infection and clinical malaria during the following transmission season. Consistent with this, studies in seasonal malaria transmission areas where the dry season lasts for ∼ 6-8 months showed that asymptomatic *P. falciparum* infections at the end of dry season are associated with the increase risk of the infection [16,36] or clinical malaria during the following transmission season [14–17,37]. The main hypothesis is that although individuals were treated at the end of the dry season, they continued to reside in the same environment where they were initially infected, with potentially higher risk of being re-infected. Nevertheless, individuals testing negative at the end of the dry season and being able to control new infections without developing symptoms suggests a fairly effective immunity and the ability to suppress parasites below detection levels. However, our data differs from other reports which show that asymptomatic *P. falciparum* infections at the end of the dry season were associated with a reduced risk of clinical malaria in the following transmission season [11–13,36,38]. Chronic infections and the maintenance of immunity could protect against high parasitaemia and clinical malaria [39,40]. The discrepancy between asymptomatic infections being a protection or a risk for reinfection/clinical malaria could be explained by the large range of malaria transmission intensity between the cited studies, as well as local environmental conditions. For example, a recent study conducted in The Gambia revealed that in the semi-urban villages dry season carriage was associated with an increased risk of clinical malaria during the following transmission season, whereas such risk was lower in rural villages [6]. Finally, a pooled analysis also demonstrated the importance of transmission intensity with risks of subsequent clinical malaria, as well as host age and polyclonality of infection [19] although this meta-analysis encompassed studies that evaluated clinical infections within the same asymptomatic infection.

In conclusion, this 2-years study identified village of residence, anaemia, and fever at enrolment as independent factors associated with *P. falciparum* infections in the eastern Gambia. *P. falciparum* transmission pattern seems stochastic. The data further validate the notion that asymptomatic parasitaemia carriage at the end of dry season correlates with a higher risk of infection and clinical malaria during the following transmission season. Therefore, targeting asymptomatic infections during the dry season may reduce the risk of clinical malaria in the following transmission season.

## Data Availability

All data produced in the present study are available upon reasonable request to the authors

## Acknowledgements

Our thanks go to Julia Mwesigwa and all fieldworkers involved in the study. We would like to thank the inhabitants of the four villages (Madina Samako, Njayel, Sendebu, and Karanbada) in eastern part of The Gambia who took part in this study.

## Supplementary Figures

**Supplementary Figure 1.**
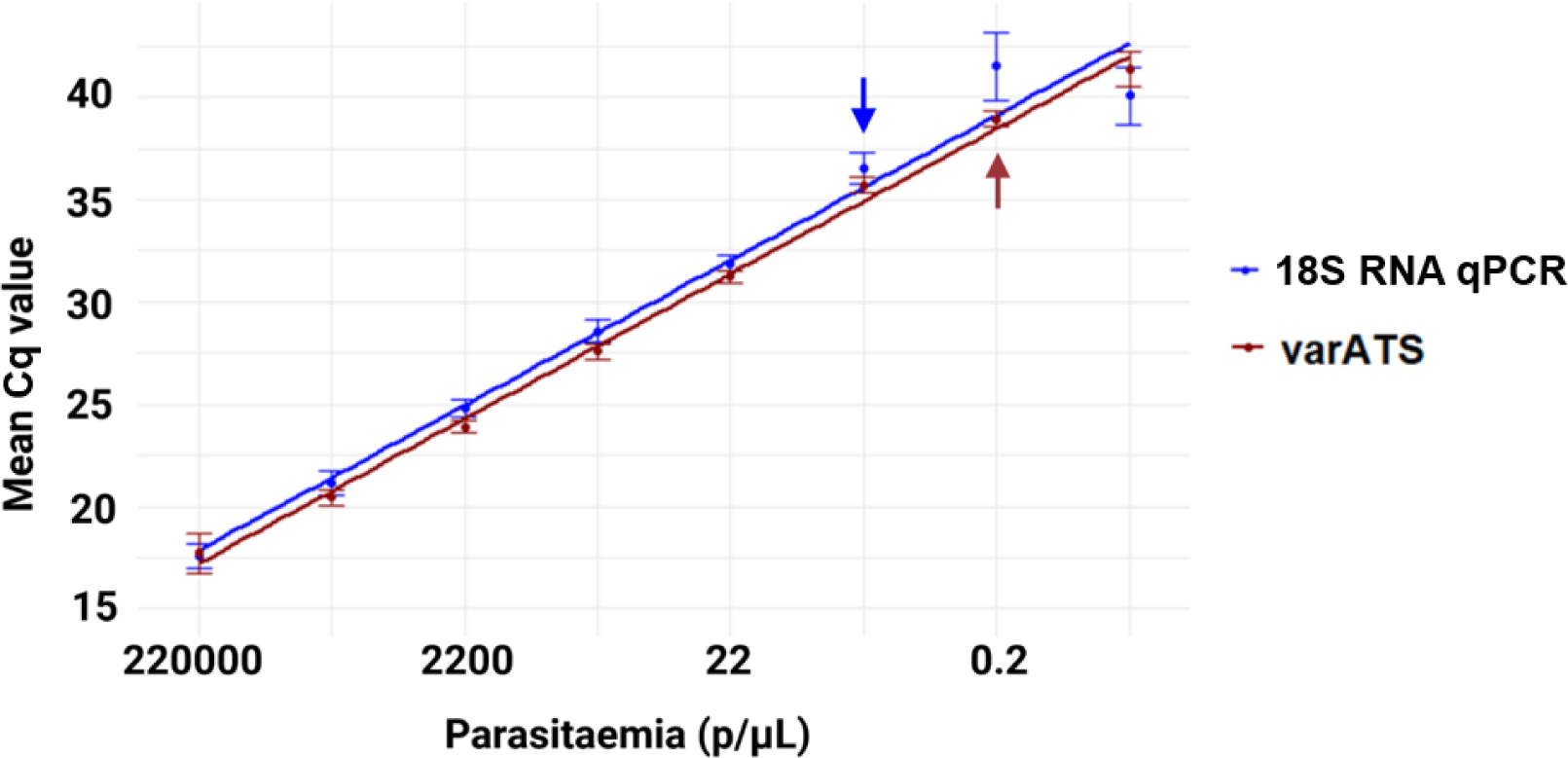
varATS and 18S RNA standard curves using 3D7 gDNA. The scatter plot represents the mean Cq value of the amplification of 8 replicates of the 10-fold serial dilution of 220000 parasites/µL of ring stage lab adapted *P. falciparum* 3D7 strain. The arrows indicate the detection limits of the two tests (2.2 parasites/µL for 18S RNA qPCR and 0.2 for varATS). Data were generated with the following method: Ring stage laboratory-adapted *P. falciparum* strain 3D7 was used to determine analytical sensitivity and parasitemia by 10-fold dilution of 220000 parasites/µL of parasitemia in 50% haematocrit from an uninfected blood donor [24]. Dilutions were prepared as a DBS and genomic DNA was extracted as described above. Each DNA sample was tested in duplicate (field isolates and 3D7 strain) and the quantification curve was made from the 3D7 standard dilution. Cut-off values for positivity for 18S rRNA qPCR and varATS were dilutions 6 (parasitaemia of 2.2 parasites/µL) and 7 (parasitaemia of 0.22 parasites/µL), respectively. Samples were defined as *P. falciparum* positive if the Ct value was less than or equal to the Ct of the cut-off. Samples that exhibited amplification for both duplicates and had a visually clear amplification curve were considered positive if their CQ value exceeded dilution 7 but remained below the upper limit of the 95% confidence interval for this parasitemia. Parasite densities were determined using standard curves generated from 3D7 genomic DNA amplifications of all dilutions of each plate with known parasite density.

**Supplementary Figure 2:**
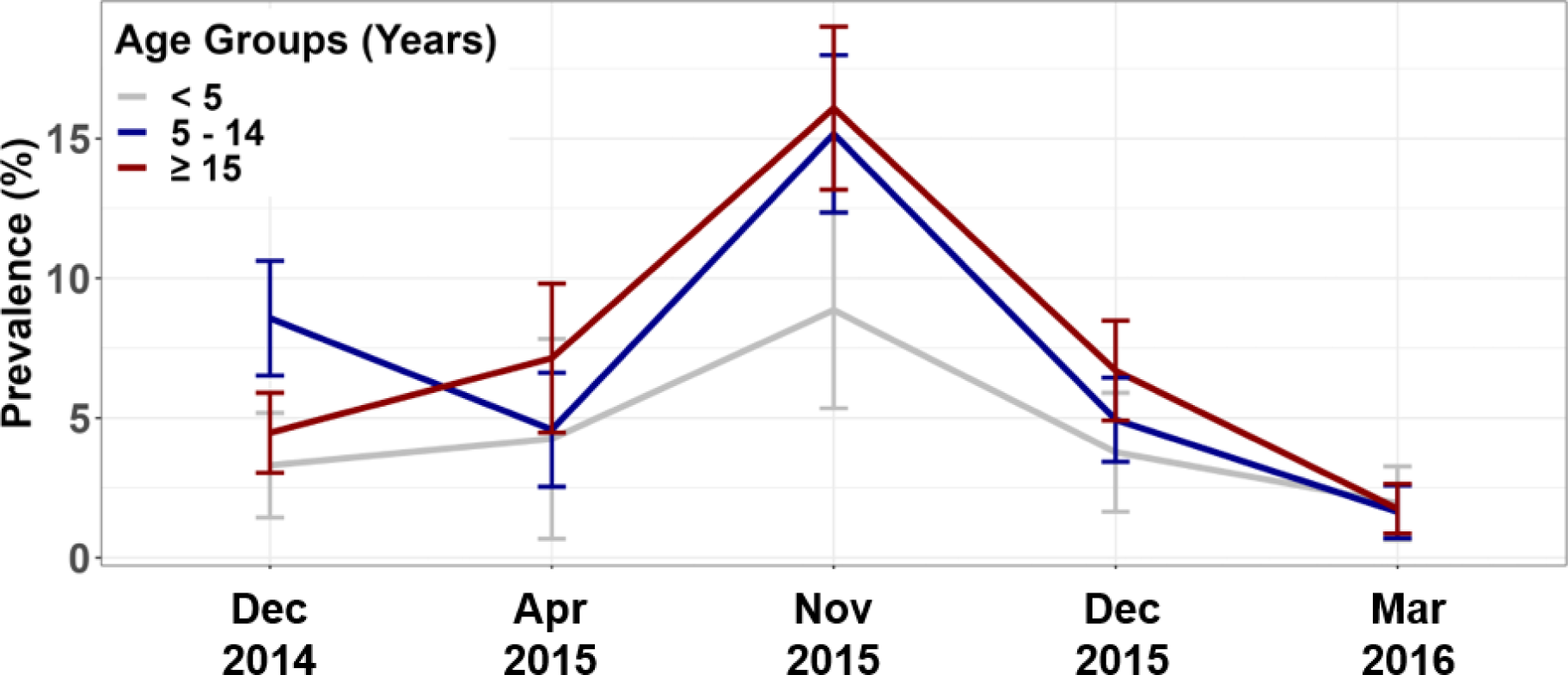
Trend analysis for *P. falciparum* infection prevalence by age groups in the first cohort, by nested PCR. Error bars represent 95% confidence intervals for prevalence.

**Supplementary Figure 3.**
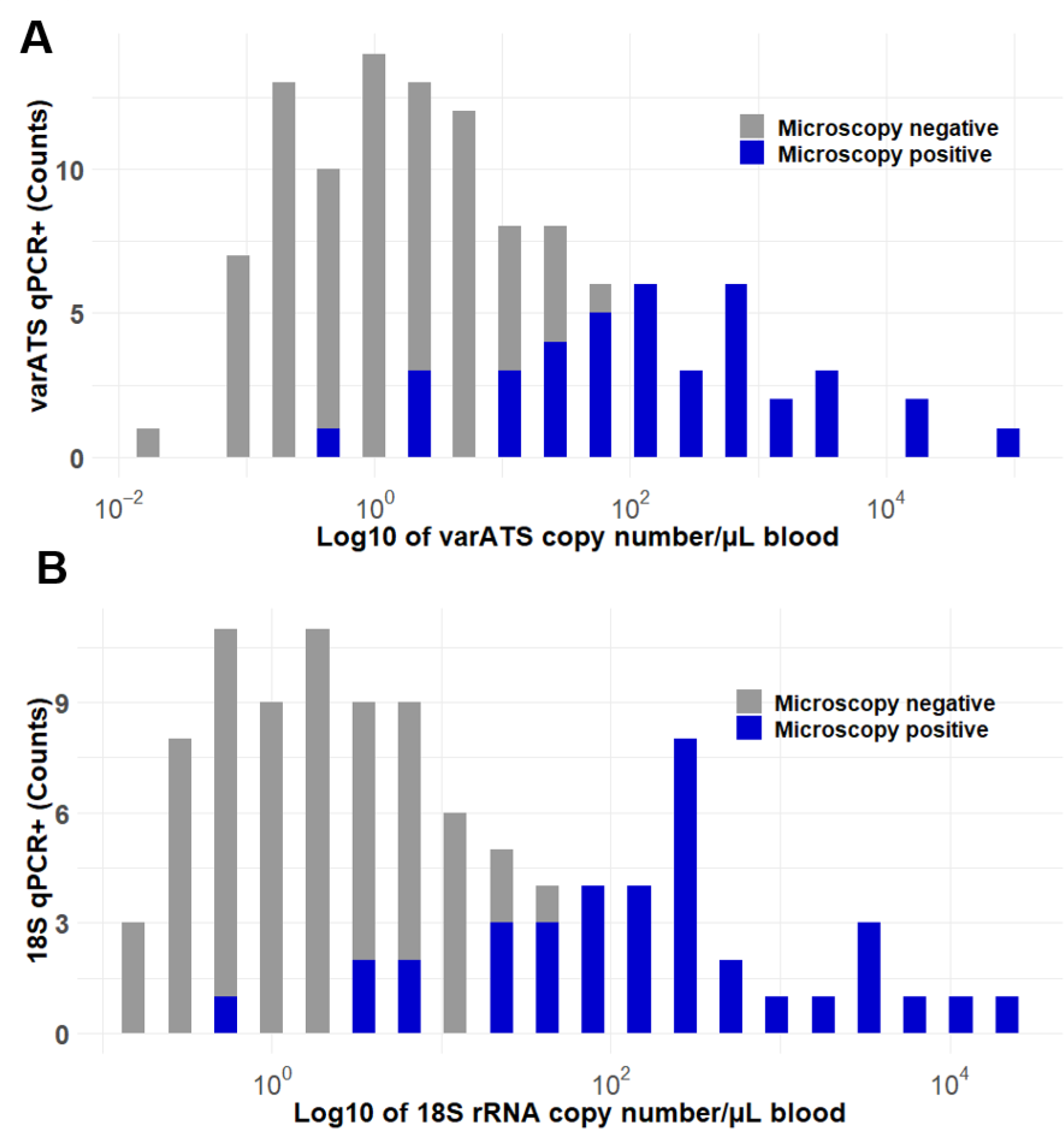
Parasites prevalence by microscopy according to parasite densities measured by varATS (A) and 18S rRNA qPCR (B) molecular tools. Parasite densities quantified by varATS and 18S rRNA qPCR are presented (log10 scale with a bin size of 20) for both microscopy negative and microscopy positive.

**Supplementary Figure 4.**
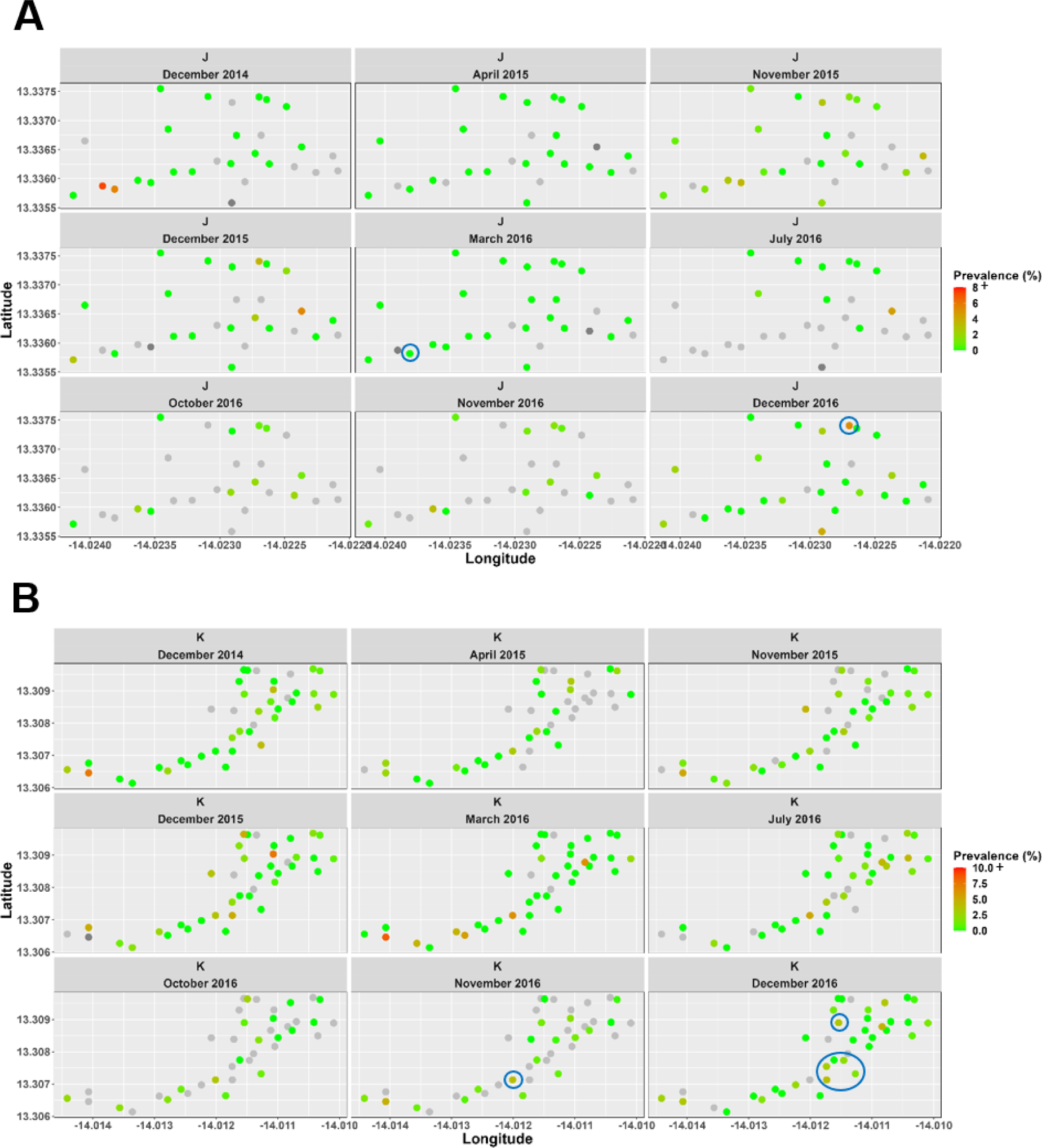
Spatio-temporal *P. falciparum* prevalence by household in villages J and K. *P. falciparum* prevalence in each household was normalized to the prevalence of infections per month in each village and only households with at least three individuals were considered. Grey colour represents the household with no data. The blue circles represent the significant hotspots (p< 0.05) determined using the Bernoulli spatial scan statistic in Satscan algorithm.

## Supplementary Tables

**Supplementary Table 1.**
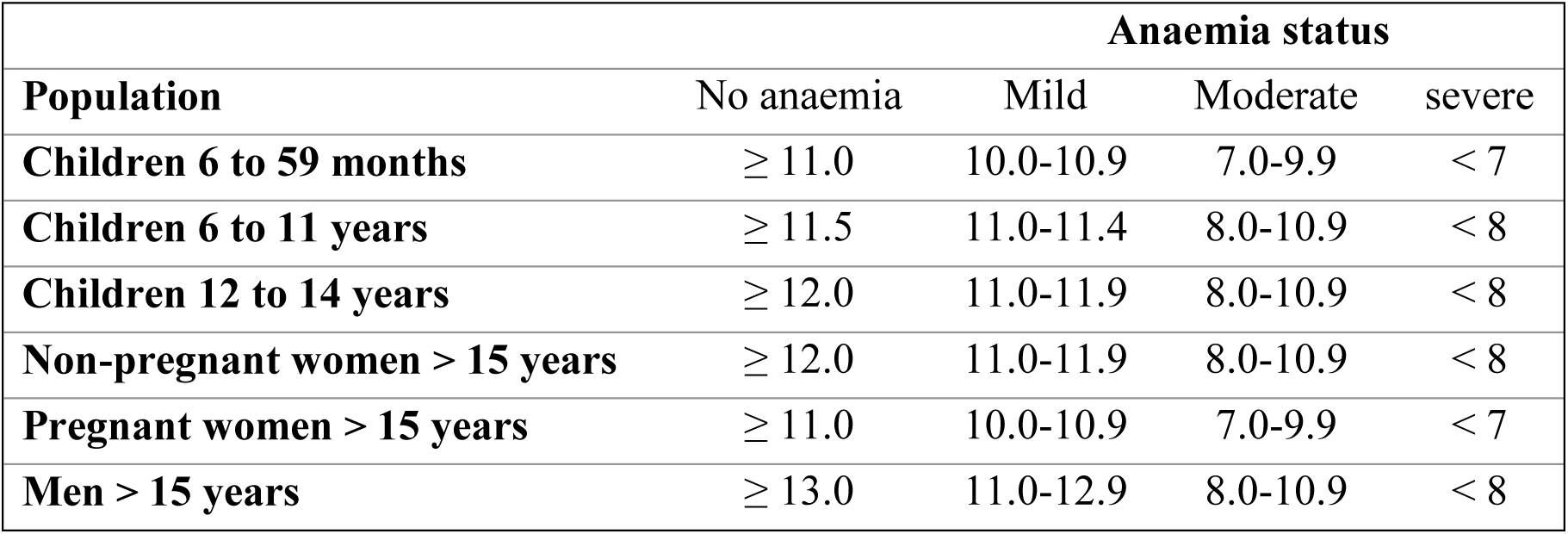
Hemoglobin levels (g/dl) and classification of anaemia (WHO, 2011)

**Supplementary Table 2:**
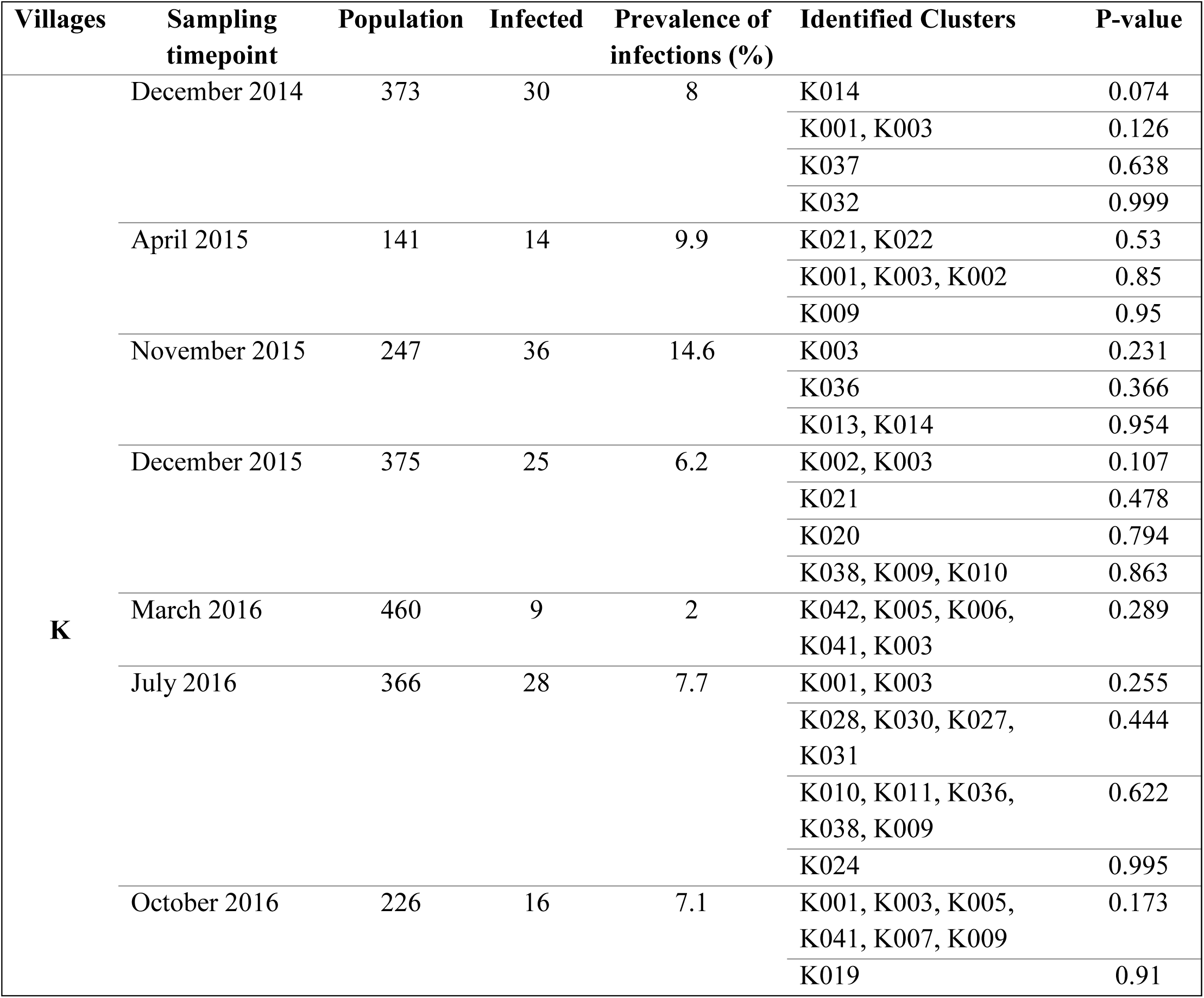
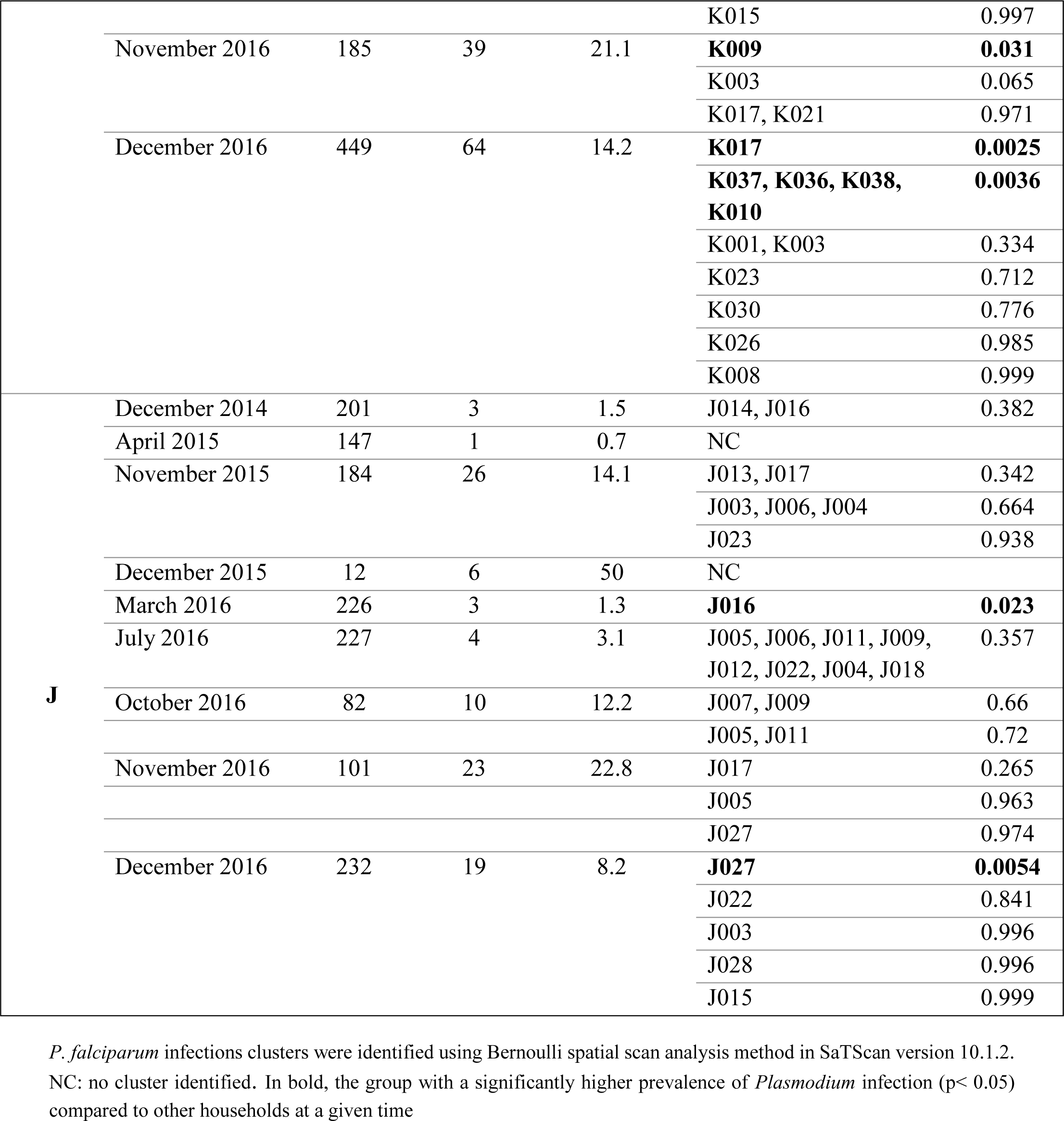
Household-based assessment of *P. falciparum* infection clusters using Bernoulli spatial scan statistic.

